# Yes, it matters: in contrast to blood plasma, serum metabolomics is confounded by platelets

**DOI:** 10.1101/2023.05.09.23289660

**Authors:** Gerhard Hagn, Samuel M. Meier-Menches, Günter Plessl-Walder, Gaurav Mitra, Thomas Mohr, Karin Preindl, Andreas Schlatter, Doreen Schmidl, Christopher Gerner, Gerhard Garhöfer, Andrea Bileck

**Author notes:** corresponding authors Dr. Andrea Bileck, Phone: Assoc. Prof. Gerhard Garhöfer, M.D.

## Abstract

Metabolomics is an emerging and powerful molecular profiling method supporting clinical investigations. For clinical metabolomics studies, serum is commonly used. Serum is collected after blood coagulation, a complex biochemical process involving active platelet metabolism. This may proof relevant as platelet counts and function may vary substantially in individuals. Applying a multi-omics analysis strategy comprising proteins and metabolites with a focus on lipid mediators, we systematically investigated serum and plasma obtained from the same healthy donors. While Biocrates MxP Quant 500 results correlated well (n=461, R^2^=0.991), lipid mediators (n=77, R^2^=0.906) and proteins (n=322, R^2^=0.860) differed substantially between serum and plasma. Actually, secretome analysis of activated platelets identified all proteins and most lipid mediators significantly enriched in serum when compared to plasma. Furthermore, a prospective, randomized, controlled parallel group metabolomics trial was performed, monitored by serum and plasma analyses. Healthy individuals received either acetylsalicylic acid, affecting platelets, or omega-3 fatty acids, hardly affecting platelets, for a period of seven days. In the acetylsalicylic acid group, serum analysis apparently demonstrated a significant drug-induced downregulation of the lipid mediators TXB2 and 12-HETE. The absence of these observation in plasma analyses suggested that these drug effects took place only during blood coagulation. Other effects of acetylsalicylic acid on alpha-linolenic acid and the fatty acid composition of triglycerides were detected both in serum and plasma. In the omega-3 fatty acid group, serum and plasma analysis results did not differ. These data strongly support the hypothesis that the serum metabolome is substantially confounded by platelets.

**Key points:**

- Serum metabolomics data are confounded by platelets
- Clinical evaluation of drug effects should be based on plasma metabolomics

## Introduction

Metabolomics represents a contemporary postgenomic analysis method for small molecules, comprising building blocks for biosynthesis, fuel for energy production along with the corresponding waste products as well as catalytically active metabolites and signaling molecules.^1^ In contrast to other biomolecules, metabolites may be formed and degraded by a variety of different and independent mechanisms, rendering data interpretation more difficult and calling for supportive machine learning algorithms.^2^ However, metabolomics data may improve the diagnosis of diseases, help to better understand disease mechanisms, and represent an important tool to practice precision medicine supporting individualized drug treatments and monitoring therapeutic outcomes.^3^

Clinical metabolomics is typically performed using serum or plasma as sample matrices. The preparation of serum implies blood coagulation before centrifuging off the particular part of full blood. In contrast, blood coagulation is inhibited on purpose by adding an anticoagulant to obtain plasma. As blood coagulation implies the functional activation of platelets, some differences in the metabolome composition of serum and plasma may be expected. However, only few studies investigated differences between serum and plasma metabolomics, reporting rather minor differences.^4-7^ A recent study provided remarkable information regarding serum and plasma protein and metabolite stability during sample preparation.^8^ However, no clear preference based on a systematic evaluation has been established up until now. With an increasing relevance of metabolomics for precision medicine, it is high time to decide for either plasma or serum resulting in improved standardization of clinical metabolomics.

Thus, we have performed systematic studies based on the assumption that platelets may affect the serum metabolome due to the blood coagulation occurring in the course of sample preparation. Detectable differences between serum and plasma were identified to be caused by platelet activation during blood coagulation. Indeed, almost all molecules significantly up-regulated in serum when compared to plasma were found to be contained in platelet releasates.

This observation raised important questions regarding the clinical investigation of drug effects by serum metabolomics. It is well established that platelet counts may vary substantially with time. In addition, all kinds of drugs interfering with inflammatory processes, arteriosclerosis or blood coagulation will affect platelet function and metabolism. This points to an unavoidable confounding of serum metabolomics data caused by the unobserved variation of platelet counts and functions. Therefore, we have conducted a prospective, randomized, controlled parallel group interventional study to assess potential differences between the serum and plasma metabolome after a seven days administration of two widely used agents, acetylsalicylic acid and omega-3 fatty acids. Acetylsalicylic acid was chosen because it is an antiphlogistic drug inhibiting enzymatic cyclooxygenase activities, thus affecting platelet activation.^9,10^ Omega-3 fatty acid supplements were chosen as, to the best of our knowledge, this should hardly affect platelet functions.

The commercial validated Biocrates MxP Quant 500 kit^11^ was used for serum and plasma metabolomics. In parallel, proteome profiling was performed to support data interpretation. Furthermore, fatty acids and lipid mediators were analyzed with an eicosadomics assay established in our laboratory, as platelets are actively involved in the formation of these special class of metabolites known to be involved in many diseases.^12^ The prospective design under tightly controlled conditions was chosen to make potential differences in the metabolomics outcomes, dependent on the choice for serum or plasma obtained from the same individuals, fully transparent and understandable.

## Methods

### Study Design and Subjects

Subjects were recruited by the Department of Clinical Pharmacology at the Medical University of Vienna. The study protocol was approved by the Ethics Committee of the Medical University of Vienna (EC No.: 2250/2020) and the Austrian competent authorities. The study was conducted in concordance with the Declaration of Helsinki and Good Clinical Practice (GCP) guidelines of the European Union. Written informed consent was obtained from all study participants prior to study entry. The study design was a randomized, controlled parallel group study.

Subjects were only included if no abnormalities were found at the screening visit. Exclusion criteria compiled symptoms of a clinically relevant illness in the 3 weeks before the first study day, a severe medical condition as judged by the investigator or the usage of any concomitant medication (except contraceptives) or dietary supplements within three weeks before the first study day.

### Randomization and study medication

Subjects were randomized to receive either acetylsalicylic acid or Omega 3 capsules for 7 days. One study cohort was instructed to take 500 mg acetylsalicylic acid (Aspirin® 500 mg acetylsalicylic acid, Cellulose powder, maize starche) per day in the evening whereas the second study cohort was instructed to take two Omega 3 complex 870 mg capsules (Br. Böhm Omega 3 capsules, 1017 mg cold water fish oil equivalent to 870 mg Omega-3, consisting of 420 mg EPA, 330 mg DHA, 5μg Vitamin D equivalent to 200 IU, 6 mg Vitamin E, 30 mg Co-enzyme Q10) per day in the evening.

### Sample collection and processing

Blood samples from the subjects were obtained at baseline (study day 1) and after 7 days intake of the study medication (study day 2). On both study days two blood samples (one for serum and one for plasma analysis) were obtained from each subject. Regarding EDTA-anticoagulated plasma sample, 6 mL K3EDTA tubes (Vacuette, Greiner Bio-One GmbH, Kremsmünster, Austria) were carefully inverted two times after blood draw and then centrifuged immediately at room temperature at 2000 g for 10 min. Then, 500 µL of plasma were transferred into pre-labelled Eppendorf safe-lock tubes. In contrast, filled 6 mL serum collection tubes (Vacuette, Greiner Bio-One GmbH, Kremsmünster, Austria) were carefully inverted after the blood draw to mix the blood and the clotting activator and placed to sit upright for 15 to 30 minutes to allow the clot to form. After clot formation, tubes were centrifuged at room temperature at 2000 g for 10 min. Again, 500 µL of serum were transferred into pre-labelled Eppendorf safe-lock tubes All samples were stored at -80°C until analysis.

### Serum and plasma proteomics

Serum and EDTA-anticoagulated plasma samples were diluted 1:20 in lysis buffer (8 M urea, 50 mM TEAB, 5 % SDS), heated at 95 °C for 5 min prior to determining the protein concentration using a BCA assay. Enzymatic digest of 20 µg of protein sample was achieved by applying the ProtiFi S-trap technology.^13^ In short, solubilized protein was reduced and carbamidomethylated using 64 mM dithiothreitol (DTT) and 48 mM iodoacetamide (IAA), respectively. Trapping buffer (90 % v/v methanol, 0.1M triethylammonium bicarbonate) was added to each sample before loading them onto the S-trap mini cartridges. Afterwards, samples were washed and subsequently digested using Trypsin/Lys-C Mix at 37 °C for 2 hours. Peptides were then eluted, dried and stored at -20 °C until LC-MS/MS analyses.

LC-MS/MS analysis was performed as described previously.^12,14,15^ Briefly, Dried peptide samples were reconstituted by adding 5 µL of 30 % formic acid (FA) containing four synthetic standard peptides and subsequent dilution with 40 µL of loading solvent (97.9 % H2O, 2 % ACN, 0.05 % trifluoroacetic acid). Thereof, 1 µL was injected into the Dionex Ultimate3000 nanoLC-system (Thermo Fisher Scientific). Pre-concentration of injected peptides was accomplished on a pre-column (2 cm□×□75 µm C18 Pepmap100; Thermo Fisher Scientific) run at a flow rate of 10 µL/min using mobile phase A (99.9 % H2O, 0.1 % FA). Subsequent peptide separation was achieved on an analytical column (25 cm x 75 µm, 1.6 µm C18 Aurora Series emitter column (Ionopticks)) by applying a flow rate of 300 nL/min and using a gradient of 7 % to 40 % mobile phase B (79.9 % ACN, 20 % H2O, 0.1 % FA) over 43 min, resulting in a total LC run time of 85 min including washing and equilibration steps. Mass spectra of separated peptides were generated using the timsTOF Pro mass spectrometer (Bruker) equipped with a captive spray ion source run at 1650 V. Further, the mass spectrometer was operated in the Parallel Accumulation-Serial Fragmentation (PASEF) mode and a moderate MS data reduction was applied. The m/z scan range was set from 100 to 1700 to record MS and MS/MS spectra and the 1/k0 scan range from 0.60 – 1.60 V.s/cm2 with a ramp time of 100 ms to achieve trapped ion mobility separation. All experiments were performed with 10 PASEF MS/MS scans per cycle leading to a total cycle time of 1.16 s. Furthermore, the collision energy was ramped as a function of increasing ion mobility from 20 to 59 eV and the quadrupole isolation width was set to 2 Th for m/z < 700 and 3 Th for m/z > 700.

LC-MS/MS data analysis including protein identification and label-free quantification (LFQ) was accomplished using the publicly available software package MaxQuant 1.6.17.0 running the Andromeda search engine.^16^ Therefore, raw data were searched against the SwissProt database “homo sapiens” (version 141219 with 20380 entries) including an allowed peptide tolerance of 20 ppm, a maximum of two missed cleavages, carbamidomethylation on cysteins as fixed modification as well as methionine oxidation and N-terminal protein acetylation as variable modification as search parameters. A minimum of one unique peptide per protein was set as search criterium for positive identifications. Additionally, the “match between runs” option was applied, using a 0.7 min match time window and a match ion mobility window of 0.05 as well as a 20 min alignment time window and an alignment ion mobility of 1. For all peptide and protein identification a false discovery rate (FDR)≤0.01 was set. Using the Perseus software^17^ (version 1.6.14.0) identified proteins were filtered for reversed sequences as well as common contaminants and annotated according to the different study groups. Then, LFQ intensity values were transformed (log2(x)), and proteins were additionally filtered for their number of independent identifications (protein has to be identified in 70% of the samples corresponding to at least one group). Finally, missing values were replaced from a normal distribution (width: 0.3; down shift: 1.8) and a principal component analysis (PCA) was performed.

### LC-MS/MS analysis of fatty acids and lipid mediators in serum and plasma

LC-MS/MS analysis of fatty acids and derivatives was performed as described previously.^12,15,18^ Briefly, frozen serum and EDTA-anticoagulated plasma samples were freshly thawed on ice. Thereof, 400 µL was mixed with cold EtOH (1.6 mL, abs. 99%, -20°C; AustroAlco) including an internal standard mixture of 12S-HETE-d8, 15S-HETE-d8, 5-Oxo-ETE-d7, 11,12-DiHETrE-d11, PGE2-d4 and 20-HETE-d6 (each 100 nM; Cayman Europe, Tallinn, Estonia) to achieve protein precipitation and stored at -20°C over-night. After centrifugation (30 min, 4536 g, 4°C), supernatants were transferred into new 15 mL Falcon^TM^ tubes and EtOH was evaporated *via* vacuum centrifugation at 37°C until the original sample volume (400 µL) was restored. Solid phase extraction (SPE) was performed by loading samples onto preconditioned StrataX SPE columns (30 mg mL−1; Phenomenex, Torrance, CA, USA) using Pasteur pipettes. After washing with 5 mL of MS grade water samples were eluted with ice-cold MeOH (500 µL; MeOH abs.; VWR International, Vienna, Austria) containing 2 % formic acid (FA; Sigma-Aldrich). MeOH was evaporated under a gentle stream of nitrogen at room temperature. Dried samples were then reconstituted in 150 µL reconstitution buffer (H2O:ACN:MeOH + 0.2 % FA–vol% 65:31.5:3.5) and subsequently measured via LC-MS/MS.

For LC-MS/MS analyses, analytes were first separated using a Thermo Scientific^TM^ Vanquish^TM^ (UHPLC) system equipped with a Kinetex® C18 column (2.6 μm C18 100 Å, LC Column 150 × 2.1 mm; Phenomenex®) applying a gradient flow profile (mobile phase A: H2O + 0.2% FA, mobile phase B: ACN:MeOH (vol% 90:10) + 0.2% FA) starting at 35% B and increasing to 90% B (1–10 min), further increasing to 99% B within 0.5 min and held for 5 min. Solvent B was then decreased to the initial level of 35% within 0.5 min and the column was equilibrated for 4 min, resulting in a total run time of 20 min per sample. The flow rate was kept at 200 μL min−1 and the column oven temperature at 40°C. Twenty µL of each sample was injected and all samples were analyzed in technical duplicates. Subsequent mass spectrometric analyses were accomplished on a Q Exactive^TM^ HF orbitrap high-resolution mass spectrometer (Thermo Fisher Scientific, Austria), equipped with a HESI source for negative ionization. The MS scan range was set to 250-700 m/z with a resolution of 60,000 (at m/z 200) on the MS1 level. A Top 2 method was applied for fragmentation (HCD 24 normalized collision energy), as well as an inclusion list covering 33 m/z values specific for well-known eicosanoids and precursor molecules. The resulting fragments were analyzed on the MS2 level at a resolution of 15,000 (at m/z 200). Operating in negative ionization mode, a spray voltage of 3.5 kV and a capillary temperature of 253 °C were applied. Sheath gas was set to 46 and the auxiliary gas to 10 (arbitrary units).

For subsequent data analysis, generated raw files were checked manually using Thermo Xcalibur^TM^ 4.1.31.9 (Qual browser) and compared with reference spectra from the Lipid Maps depository library from July 2018.^19^ Peak integration was performed using the TraceFinder^TM^ software package (version 4.1—Thermo Scientific, Vienna, Austria). The resulting data were loaded into the R software package environment (version 4.2.0)^20^ and peak areas of each analyte were log2-transformed. For normalization, the mean peak area of the internal standards was subtracted from the analyte peak areas to correct for variances arising from sample extraction and LC-MS/MS analysis. Then, each log2-transformed area was increased by adding (x+20) to obtain a similar value distribution compared to label-free quantification in proteomics. Missing values were imputed using the minProb function of the imputeLCMD package (version 2.1).^21^ Principal component analysis was performed using the Perseus software (version 1.6.14.0).^17^

### Serum and plasma metabolomics

Targeted metabolomics experiments of serum and plasma were conducted by applying the MxP® Quant 500 Kit (Biocrates Life Sciences AG, Innsbruck, Austria) as described previously.^12,15^ Therefore, 10 µL of sample was used and the kit was performed according to the manufacturer’s instructions. Measurements were carried out using LC-MS/MS and flow injection (FIA)-MS/MS analyses on a Sciex 6500+ series mass spectrometer coupled to an ExionLC AD chromatography system (AB Sciex, Framingham, MA, USA), utilizing the Analyst 1.7.1 software with hotfix 1 (also AB SCIEX). All required standards, quality controls and eluents were included in the kit, as well as the chromatographic column for the LC-MS/MS analysis part. Phenyl isothiocyanate (Sigma-Aldrich, St. Louis, USA) was purchased separately and was used for derivatization of amino acids and biogenic amines according to the kit manual. Preparation of the measurement worklist as well as data validation and evaluation were performed with the software supplied with the kit (MetIDQ-Oxygen-DB110-3005, Biocrates Life Sciences). Out of the 630 included analytes, a total of 461 metabolites showed signal intensities within the quantification window and were further evaluated. Principal component analysis was performed using the Perseus software (version 1.6.14.0).^17^

### Platelet isolation, activation and LC-MS/MS analyses

Whole blood of six healthy donors (three male and three female) in the age range of 26 to 51 years were collected in biological duplicates with one week in between the donations. Each donor gave written consent and the study was approved by the ethics committee of the Medical University of Vienna in accordance with the Declaration of Helsinki (EC 1430/2018). No medical substances interfering with the normal physiology of platelets such as aspirin, paracetamol or ibuprofen were taken by the donors 48 hours prior to blood donation. Two CPDA (citrate-phosphate-dextrose-adenine)-S-Monovette tubes (Sarstedt) of venous blood were collected per donor and donation. To isolate platelet rich plasma (PRP), the tubes were centrifuged for 20 min at 100 g with acceleration and deceleration set to 4.

To purify platelets, size exclusion chromatography using 2 % B agarose beads (50-150 μm; abtbeads.es) was performed. Therefore, columns were equipped with a cotton frit and 20 mL of reconstituted agarose bead solution diluted 1:2 in RPMI medium (1X with L-Glutamine; Gibco, Thermo Fischer Scientific, Austria). Columns were washed with 2 mL RPMI medium before 1 ml of PRP was carefully pipetted to the column and topped with RPMI. The fraction containing purified platelets was collected and divided in two aliquots, one for platelet activation and one serving as control. To achieve platelet activation, ionomycin calcium salt (Sigma-Aldrich) was added to one aliquot to a final concentration of 1 μM. All samples were incubated for 15 min at room temperature before centrifugation at 2000 g for 5 min. The supernatant was transferred into new tubes and protein precipitation was performed by adding ice cold ethanol (LC-MS grade) in a ratio of 1:5. Additionally, 5 μL of an internal standard mixture of 12S-HETE-d8, 15S-HETE-d8, 5-Oxo-ETE-d7, 11,12-DiHETrE-d11, PGE2-d4 and 20-HETE-d6 (each 100 nM; Cayman Europe, Tallinn, Estonia) were added to each sample. Samples were then stored at -20°C. After overnight precipitation, samples were centrifuged for 30 min at 4536 g at +4°C. The supernatant was then transferred into new 15 mL Falcon^TM^ tubes and submitted to the lipid extraction workflow in order to enable LC-MS/MS analysis of fatty acids and lipid mediators as described above for serum and plasma samples. The remaining pellets representing secreted proteins were dried in an exsiccator and submitted to the proteomics workflow as described above for serum and plasma samples.

### Statistical analysis and graphical visualization

For statistical analyses log2 transformed expression values were loaded into R,^22^ and fitted to a linear model using LIMMA^23^ with subjectID as pairing variable where appropriate. P-values were adjusted for multiple testing according to Benjamini-Hochberg.^24^ Volcano plots representing log2 fold-changes on the x-axis and -log adjusted p-values on the y-axis were generated using GraphPad Prism Version 6.07 (2015). Molecules displaying a fold-change of ≥ or ≤ 2 and an adjusted p-value of ≤ 0.05 were considered as statistically significant. Pearson r correlation analyses between serum and plasma were performed separately for proteins, fatty acid and lipid mediators as well as metabolites using GraphPad Prism Version 6.07 (2015). The effect of Aspirin on a subset of 136 triglycerides (TGs) was shown by means of a linear regression and Spearman r correlation analyses, again using GraphPad Prism Version 6.07 (2015). Therefore, fold-changes of C:16 and C:18 TGs in serum and plasma, respectively, before and after 7 days of Aspirin intake were correlated with the sum of C-atoms of the two remaining fatty acids. Heatmap visualizing fold-changes of C:16 and C:18 TGs in serum and plasma, respectively, before and after 7 days of Aspirin intake was generated using Microsoft Excel.

### Data Sharing Statement

All proteomics data was submitted to the ProteomeXchange Consortium (http://proteomecentral.proteomexchange.org) and is available in the PRIDE partner repository^25^ with the dataset identifier PXD041781 and PXD041785.

Data derived from all metabolomics as well as fatty acid and lipid mediators’ analyses produced in the present study are available upon request to the authors.

## Results

### Molecular characterization of serum and plasma samples

In order to investigate the molecular composition of plasma in comparison to serum of untreated healthy donors, venous blood was drawn from six individuals. Serum and plasma were obtained from each blood donation. Proteome profiling based on label-free shotgun analysis identified 322 proteins in total and demonstrated the almost complete loss of the fibrinogen subunits in serum, as expected due to blood coagulation. Seven proteins were apparently upregulated in serum when compared to plasma (Figure 1A, Supplementary Table S1). Principle component analysis (PCA) separated serum and plasma samples (Figure 1B). The correlation coefficient of proteins detected in serum and plasma was found to be R^2^=0.860 (Figure 1C). The eicosadomics assay, comprising 77 different molecules, identified 9 molecules significantly different between serum and plasma (Figure 1D, Supplementary Table S1). The PCA hardly separated serum and plasma samples (Figure 1E) and the correlation coefficient was found to be R^2^=0.906 (Figure 1F). The metabolomics kit, comprising a total of 461 molecules, identified only 4 metabolites with significant concentration differences between serum and plasma (Figure 1G). Here, the PCA did not separate serum and plasma samples (Figure 1H) and the correlation coefficient was found as high as R^2^=0.991 (Figure 1I, Supplementary Table S1). This data suggested only minor differences in the metabolome as determined from serum and plasma isolated from healthy donors.

**Figure 1:**
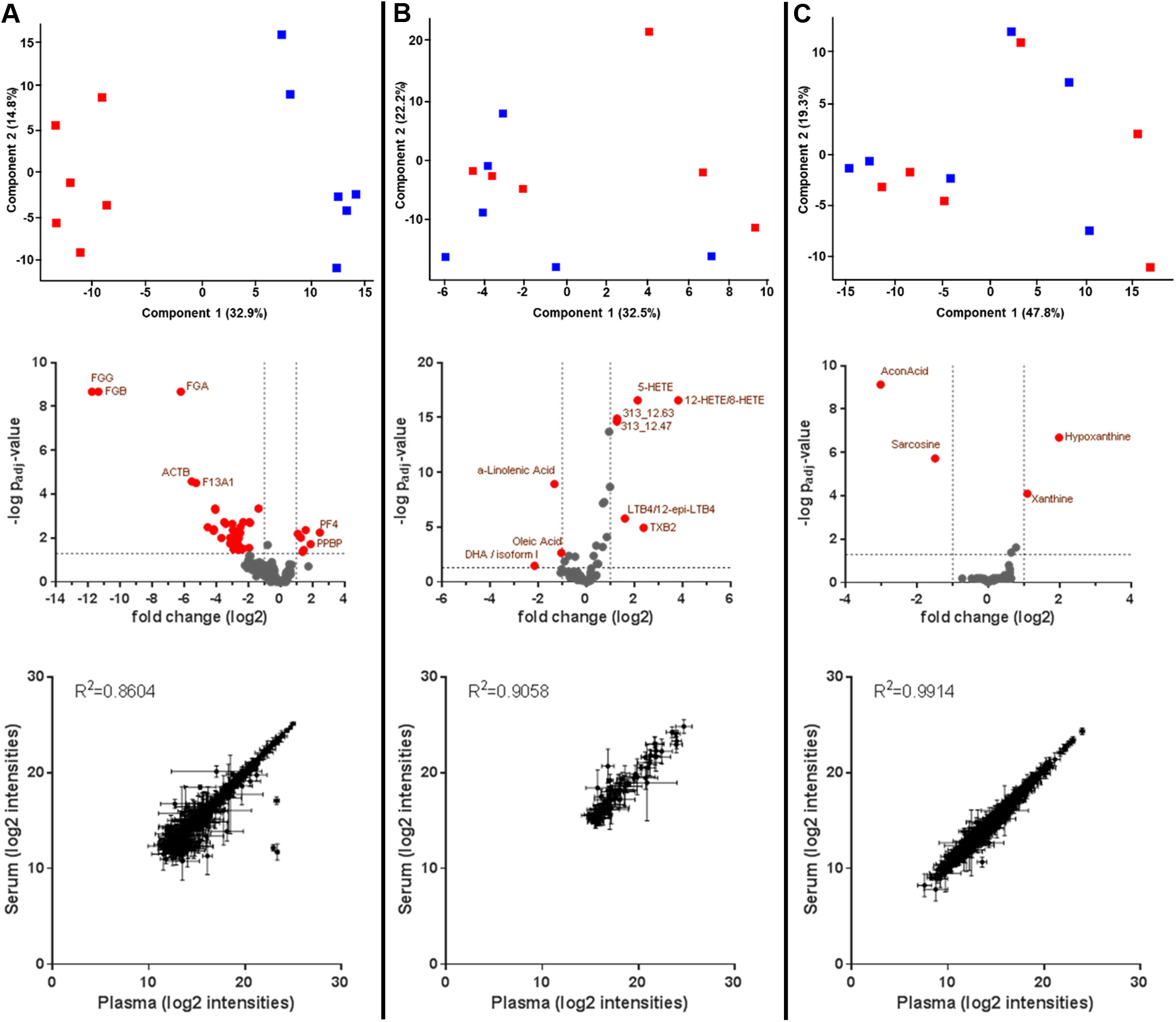
Molecular characterization of serum and plasma samples. Principal component analyses (red = serum samples; blue = plasma samples), volcano plots showing significant differences of molecules between serum and plasma (positive fold-changes mean higher abundance in serum compared to plasma) as well as correlation analyses are shown for (A) proteins, (B) fatty acids and lipid mediators and (C) metabolites. For statistical analyses, data were fitted to a linear model using LIMMA with subjectID as pairing variable and p-values were adjusted for multiple testing according to Benjamini-Hochberg. Molecules displaying a fold-change of ≥ or ≤ 2 and an adjusted p-value of ≤ 0.05 were considered as statistically significant and marked in red in the corresponding volcano plot.

### Main differences between serum and plasma are a consequence of platelet activation

A functional annotation of the proteins apparently upregulated in serum, when compared to plasma, using the DAVID Bioinformatics Resources^26,27^ suggested platelets as their potential origin (GOBP: platelet activation, Benjamini-Hochberg adjusted p-value = 3.5E-3; GOCC: platelet alpha granule lumen, Benjamini-Hochberg adjusted p-value = 9.7E-9). In order to investigate this hypothesis in more detail, platelets were isolated and activated *in vitro* by the addition of ionomycin. The following platelet aggregation was found to be accompanied by the release of 786 proteins in the supernatant and the release of 20 fatty acids and lipid mediators (Figure 2, Supplementary Table S2). Remarkably, all proteins (other than antibodies) with increased abundance values in serum, when compared to plasma, were also found to be released from platelets upon aggregation. Similarly, most eicosanoids with high abundance values in serum were found to be released from platelets upon aggregation (Table 1). These observations substantiated the hypothesis that platelet releasates, formed during blood coagulation, contributed substantially to the molecular composition of serum.

**Figure 2:**
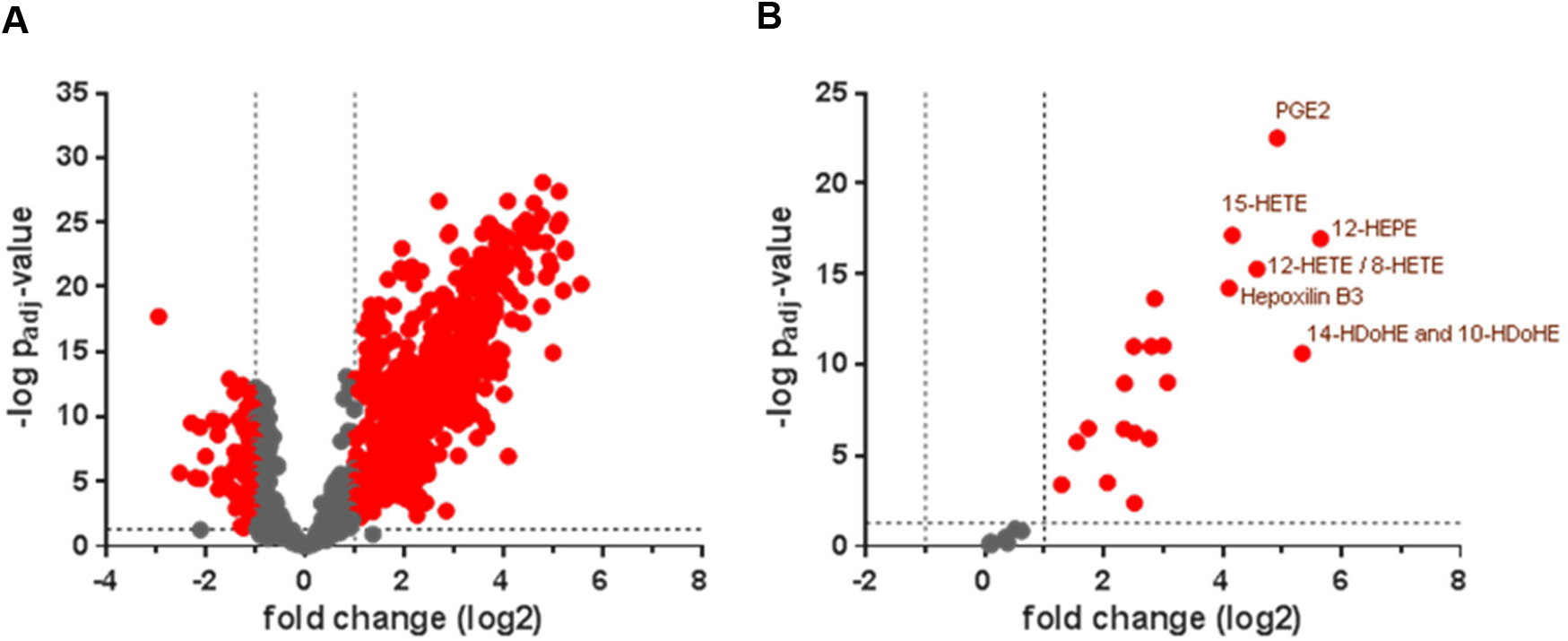
Molecular characterization of platelet releasates upon activation. Volcano plots showing significant regulations of (A) proteins and (B) lipid mediators upon platelet activation. P-values were calculated based on a linear model using LIMMA with subjectID as pairing variable and adjusted for multiple testing according to Benjamini-Hochberg. Molecules displaying a fold-change of ≥ or ≤ 2 and an adjusted p-value of ≤ 0.05 were considered as statistically significant and marked in red.

**Table 1:** Proteins and lipid mediators differing serum and plasma are derived from platelets. Log2 fold-changes of molecules as well as Benjamini-Hochberg adjusted p-values of significantly upregulated molecules in serum, when compared to plasma, are shown. For each molecule it is indicated whether it was found in the supernatant of platelets and whether it was significant upon activation.

| Proteins | log2 fold-change serum vs plasma | adjusted p-value | found in supernatant of platelets | significant upon platelet activation |
| --- | --- | --- | --- | --- |
| PF4; Platelet factor 4 | 2,47 | 5,57E-03 | + |  |
| PPBP; Platelet basic protein | 1,88 | 1,91E-02 | + |  |
| VWF; von Willebrand factor | 1,46 | 3,50E-02 | + |  |
| MMRN1; Multimerin-1 | 1,39 | 4,23E-02 | + |  |
| PF4V1; Platelet factor 4 variant | 1,28 | 9,61E-03 | + |  |
| FN1; Fibronectin | 1,08 | 6,35E-03 | + |  |
| **Lipid mediators** |  |  |  |  |
| 12-HETE/8-HETE | 3,83 | 2,73E-17 | + | + |
| TXB2 | 2,39 | 1,15E-05 | + | + |
| 5-HETE | 2,14 | 2,73E-17 |  |  |
| LTB4/12-epi-LTB4 | 1,61 | 1,63E-06 |  |  |
| 313_12.63 | 1,28 | 1,28E-15 |  |  |
| 313_12.47 | 1,28 | 2,42E-15 |  |  |

### Metabolic alterations in response to acetylsalicylic acid administration apparently differ between plasma and serum

After comparing background levels of biomolecules between plasma and serum, we investigated molecular alterations in response to acetylsalicylic acid administration in a prospective, randomized, controlled parallel group trial. Proteome profiling of plasma revealed 32 proteins significantly upregulated upon drug exposure (Figure 3A). Thirty-one of those 32 proteins were found to be released by platelets upon coagulation, pointing to acetylsalicylic acid effects on platelets as plausible explanation for this observation (Supplementary Table S3). In contrast, only one protein, fibrinogen beta chain, was found to be apparently deregulated in serum upon drug exposure (Figure 3A). Thus, serum and plasma analysis results of the effects of acetylsalicylic acid administration differed substantially.

**Figure 3:**
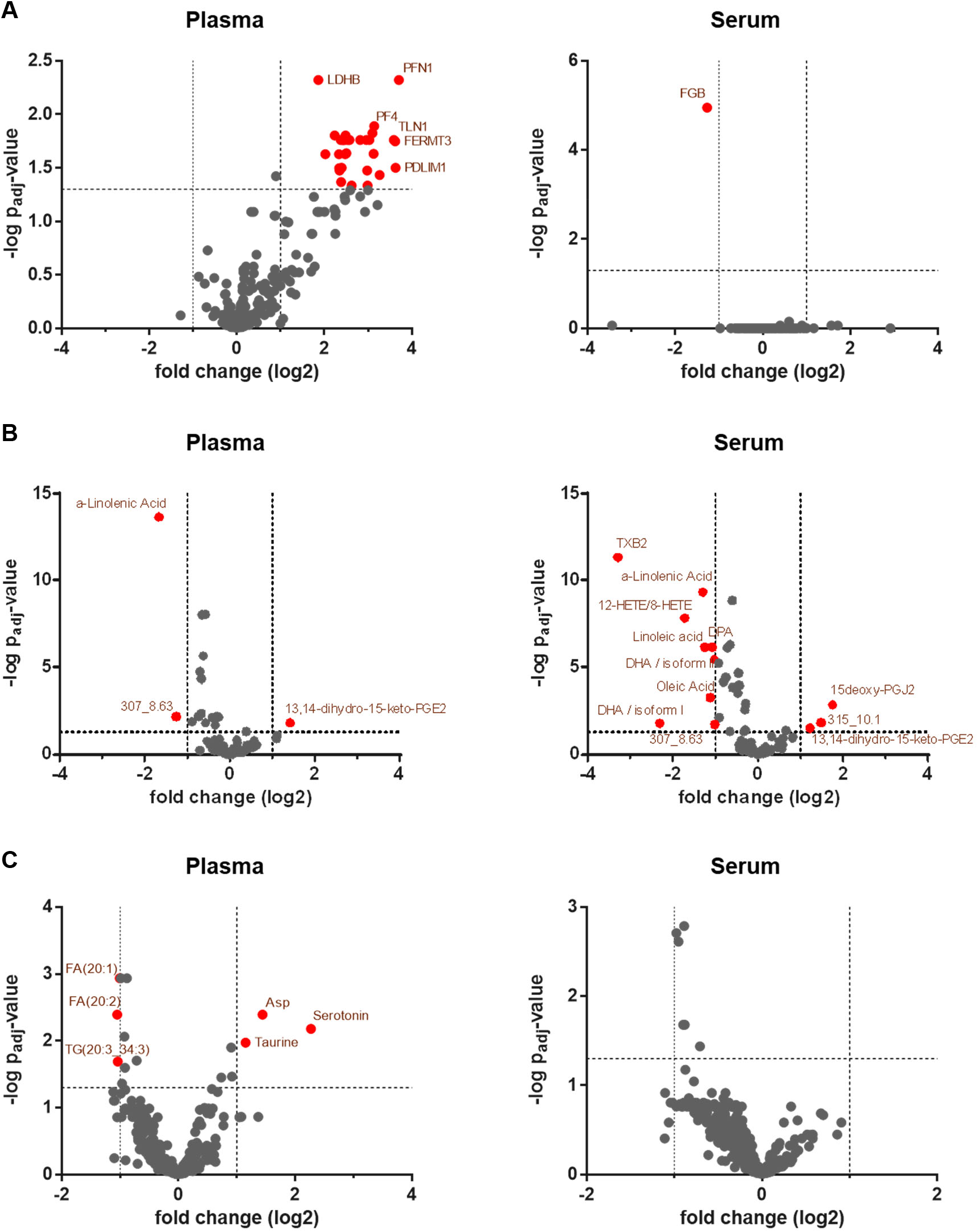
Effects of Aspirin on the molecular profile of serum and plasma, respectively. Volcano plots showing Aspirin-induced effects on A) proteins, B) lipid mediators and C) metabolites in serum and plasma, respectively. P-values were calculated based on a linear model using LIMMA with subjectID as pairing variable and adjusted for multiple testing according to Benjamini-Hochberg. Molecules displaying a fold-change of ≥ or ≤ 2 and an adjusted p-value of ≤ 0.05 were considered as statistically significant and marked in red.

The eicosadomics assay performed with the same samples demonstrated the significant downregulation of TXB2, 12-HETE and other fatty acids by acetylsalicylic acid in serum (Figure 3B, Supplementary Table S3). TXB2 and 12-HETE were among the most abundant eicosanoids released by platelets upon coagulation (Table 1, Supplementary Table S2). Thus, this data again pointed to the known mode of action of acetylsalicylic acid, inhibiting cyclooxygenases and platelet activation and thus the formation of these molecules.^10^ However, when analyzing the effects of acetylsalicylic acid administration in plasma, there results were hardly reproduced. Mainly alpha-linolenic acid, an omega-3 fatty acid, was found to be downregulated, whereas the eicosanoids TXB2 and 12-HETE were not affected (Figure 3B, Supplementary Table S3).

The administration of acetylsalicylic acid also induced alterations of some lipids (Figure 3C). Both serum and plasma analyses identified the downregulation of alpha-linolenic acid (Figure 3B) accompanying a shift in the triglyceride (TG) composition towards higher levels of TGs with shorter side chains (Figure 4). Spearman r correlation analyses between fold-changes of a subset of 136 different C:16 and C:18 TGs, before and after 7 days of acetylsalicylic acid intake, and the sum of C-atoms of the two remaining fatty acids clearly demonstrated a significant correlation in plasma and serum (Figure 4B, C), pointing to consistent drug effects *in vivo*.

**Figure 4:**
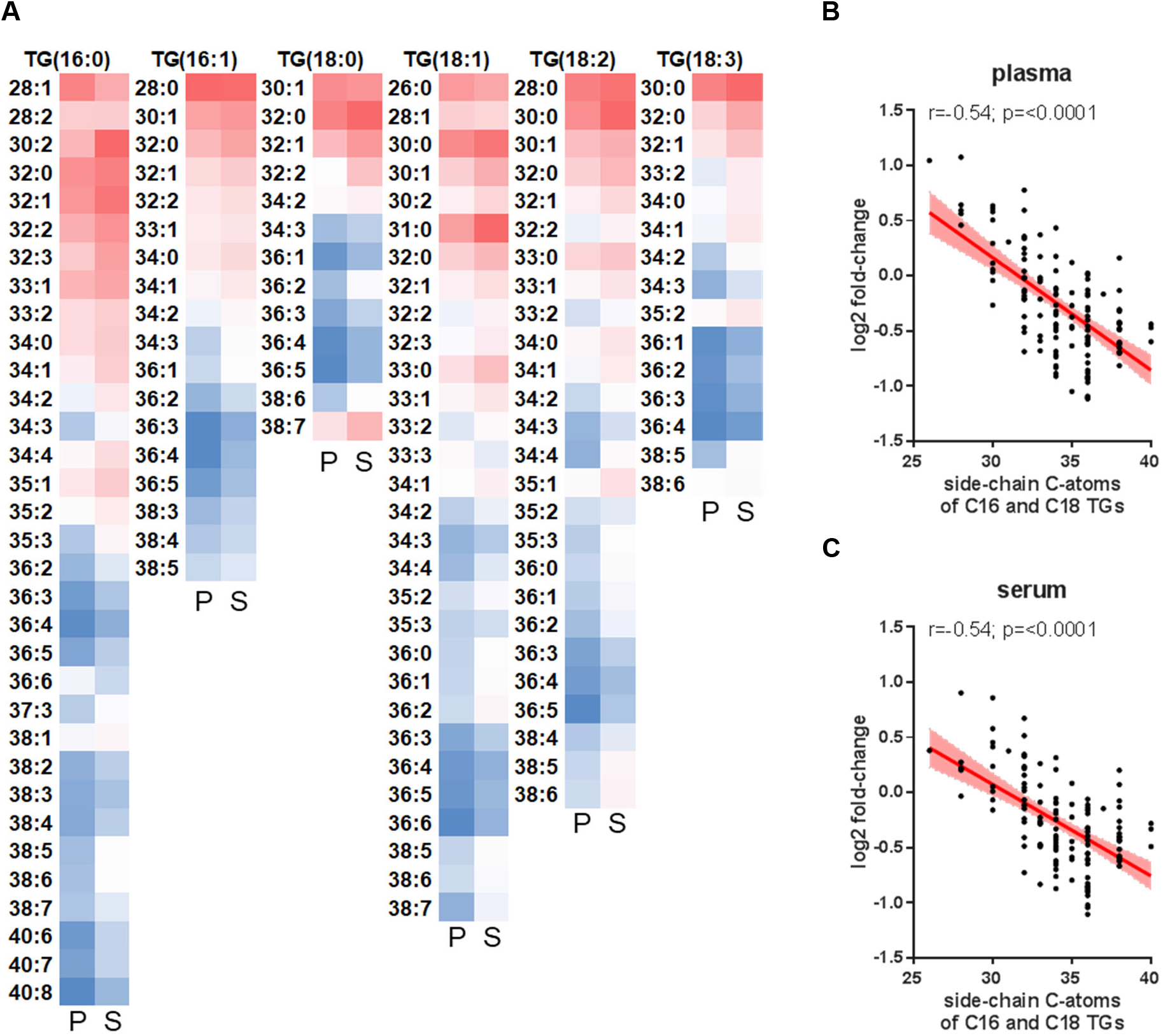
Effects of Aspirin consumption on the fatty acid composition of triglycerides. (A) Heatmap displaying the fold-changes of triglycerides (TGs) in plasma (P) and serum (S) when comparing samples before and after 7 days of Aspirin consumption. While fold-changes marked in red indicate upregulation of respective TGs upon Aspirin consumption, fold-changes marked in blue indicate higher levels of respective TGs before Aspirin intake. Linear regression and Spearman r correlation analyses between fold-changes of C:16 and C:18 TGs in (B) plasma and (C) serum, before and after 7 days of Aspirin intake and the sum of C-atoms of the two remaining fatty acids are shown.

With regard to other metabolites, serotonin, taurine and asparagine were found significantly upregulated in plasma upon acetylsalicylic acid administration, whereas these molecules were apparently not affected in serum samples.

### Metabolic alterations in response to omega-3 fatty acids administration did not differ between plasma and serum

Administration of omega-3 fatty acids, as performed in an independent prospective, randomized, controlled parallel group trial, resulted in the significant upregulation of EPA and 17(18)-DiHETE (Figure 5). Remarkably, these alterations were observed in both, serum and plasma samples in an almost identical fashion.

**Figure 5:**
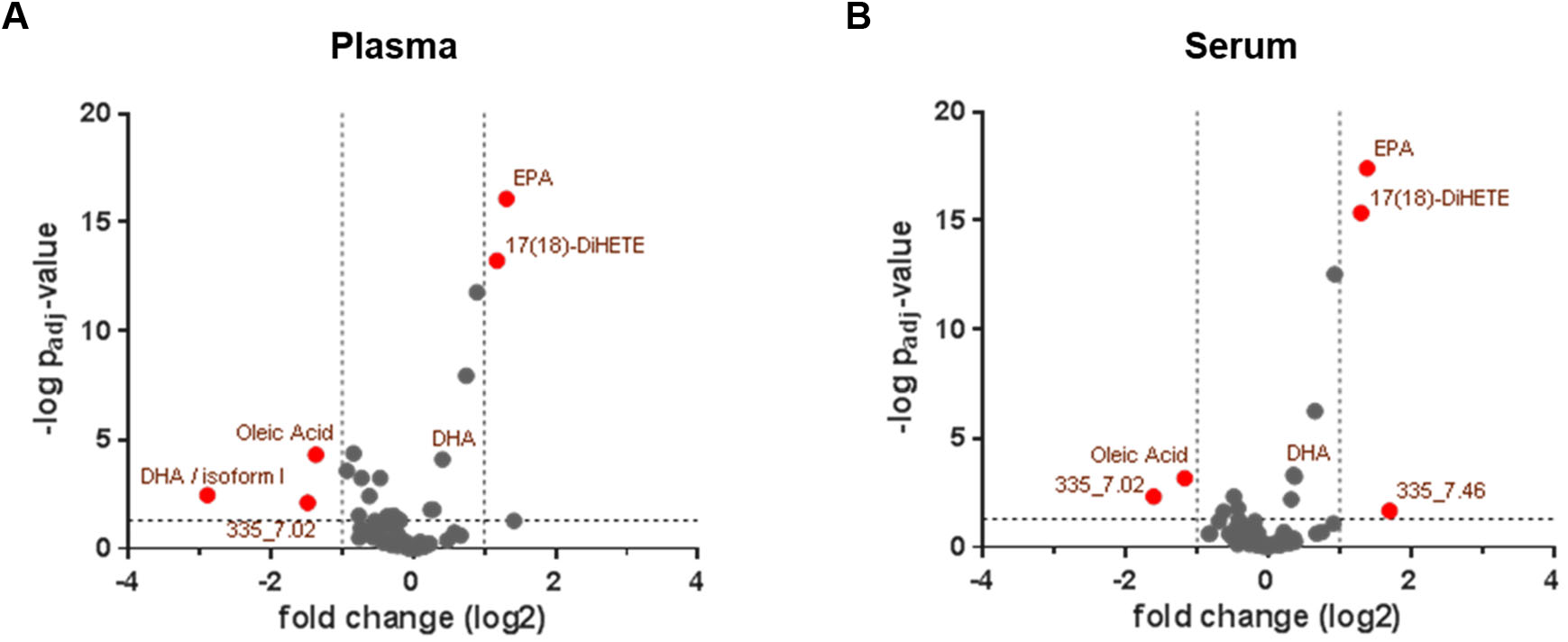
Effects of Omega-3 supplementation on fatty acids and lipid mediators in serum and plasma, respectively. Volcano plots showing Omega-3-induced effects on fatty acids and lipid mediators in (A) plasma and (B) serum. P-values were calculated based on a linear model using LIMMA with subjectID as pairing variable and adjusted for multiple testing according to Benjamini-Hochberg. Molecules displaying a fold-change of ≥ or ≤ 2 and an adjusted p-value of ≤ 0.05 were considered as statistically significant and marked in red.

### Platelets represent the main confounder responsible for differences between serum and plasma metabolomics

When investigating drug effects on the metabolism in humans, serum and plasma represent the most important sample sources. Importantly, blood coagulation specifically occurrs during serum production. The functional state of platelets *in vivo* may clearly affect molecular alterations accompanying blood coagulation (Figure 6). On the other hand, the functional state of platelets may have direct effects on the serum metabolome (Figure 6). Platelets are entities with active metabolism, upon activation they may consume, synthesize and release various metabolites. Thus, the functional state of platelets represents a confounder for the metabolome composition of serum after coagulation (Figure 6). This does not apply when analyzing plasma, as no coagulation takes place thereby.

**Figure 6:**
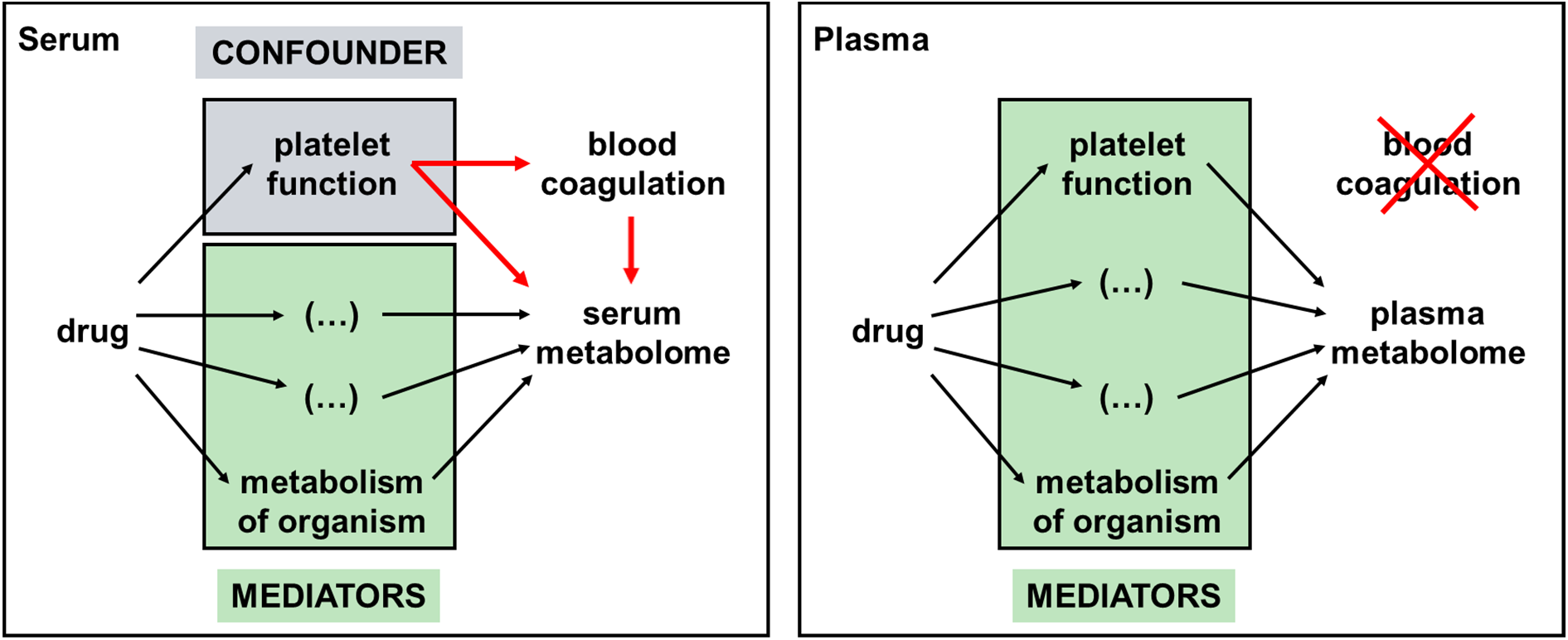
Platelets are the main confounder in serum metabolomics. The functional state of platelets *in vivo* is affecting blood coagulation during sample preparation and the serum metabolome. Thus, the functional state of platelets represents a confounder for the metabolome composition of serum. This does not apply when analyzing plasma. Here, no coagulation takes place and the functional state of platelets represents a mediator.

## Discussion

Here, we show for the first time in a controlled, randomized interventional trial in humans that serum and plasma metabolomics results may differ substantially when analyzing drug effects. The present data demonstrated that several drug-induced changes of the serum metabolome may not mirror the patientś metabolomic states but rather reflected consequences of the applied drug on platelet metabolism during blood coagulation. Thus, platelets were identified as relevant confounders for the serum metabolome, but not plasma metabolome (Figure 6).

This finding may have important practical implications. Evidently, there is a continuous rise in the number of metabolomics papers published, which are based on serum or plasma analyses. Typically, such metabolomics studies are intended to increase our current understanding for disease mechanisms and therapeutic options. However, the inherent mechanistic structure of data linking metabolic states with disease processes and understanding the consequences of drug treatment is by far not trivial. In such a complex chain of events, many important mechanisms and players may remain unobserved or unrecognized, rendering data interpretation inaccurate.

Considering important lipid mediators demonstrated that many metabolites observed to be altered when investigating the effects of acetylsalicylic acid in healthy subjects were confounded by the metabolism of platelets during blood coagulation. The data depicted in Figure 3B suggested that acetylsalicylic acid inhibited the formation of eicosanoids in the blood when analyzing serum. Understanding the mechanistic steps involved in serum sample preparation tells us this suggestion was not correct. In fact, acetylsalicylic acid inhibited eicosanoid formation during blood coagulation, occurring after blood donation. The metabolome alterations observed in plasma may thus better report actual drug effects taking place in the organism. Indeed, acetylsalicylic acid consumption seemed to have consequences beyond platelet inhibition. This was suggested by the drug-induced upregulation of serotonin, aspartic acid and taurine, in addition to an alteration in the fatty acid composition of triacylglycerols. These observations are not new, but in accordance with existing literature.^10,28-30^ However, the biological significance of these findings remains to be investigated in the future.

As the metabolomics methodology becomes more and more sensitive, we can also expect to observe more apparent disease- or drug treatment-related metabolic alterations which may be confounded and thus misleading when investigating serum samples. Plasma samples were hardly confounded, as no cellular activity was specifically induced during sample generation. Remarkably, the sample processing time from blood donation to sample freezing was apparently sufficient to cause a significant upregulation of aconitic acid in plasma when compared to serum (Figure 1C). Aconitic acid is a metabolic product formed by platelets out of citrate. This observation should remind us that also plasma collection may be accompanied by biochemical processes, requiring potential additional correction strategies for metabolomics to be established in the future.

This study highlights the implications of potential confounders on a complex data structure typically prevalent in biomedical studies. When applying metabolomics to investigate diseases and drug treatments, a direct and linear relation of a disease mechanism with a given metabolite is not to be expected. All the more a methodological standardization of metabolomics workflows is desirable. Here we suggest that plasma metabolomics may be better suitable for clinical studies than serum metabolomics as data will not be confounded by varying platelet counts and functional conditions.

## Supporting information

Supplementary Table 1 serum vs plasma

Supplementary Table 2 platelets

Supplementary Table 3 Aspirin effects

Supplementary Table 4 Omega3 eicos

## Data Availability

All proteomics data was submitted to the ProteomeXchange Consortium (http://proteomecentral.proteomexchange.org) and is available in the PRIDE partner repository with the dataset identifier PXD041781 and PXD041785.
Data derived from all metabolomics as well as fatty acid and lipid mediators analyses produced in the present study are available upon request to the authors.

## Acknowledgements

The authors are grateful to the Core Facility of Mass Spectrometry at the Faculty of Chemistry and the Joint Metabolome Facility, both members of the Vienna Life-Science Instruments (VLSI).

## Authorship Contributions

AS, DS and GG conducted clinical studies and collected samples. AB and GM performed cell culture. GPW and GM performed proteomics sample preparation. GH performed oxylipin and fatty acid analysis. SMM and KP performed metabolomics. AB, GH, SMM and KP acquired and processed data. CG, AB, GH, SMM and TM processed data. CG, AB and GG planned and supervised the study. CG and AB wrote the initial draft of the manuscript, which was edited and approved by all authors.

## Disclosure of Conflicts of Interest

The authors declare no competing financial interests.

## Notes

### Competing Interest Statement

The authors have declared no competing interest.

### Clinical Trial

NCT05775536

### Funding Statement

This study did not receive any funding

### Author Declarations

The study protocol was approved by the Ethics Committee of the Medical University of Vienna (EC No.: 2250/2020) and the Austrian competent authorities. The study was conducted in concordance with the Declaration of Helsinki and Good Clinical Practice (GCP) guidelines of the European Union.

